## Supplementary Figure 1 for "Artificial Intelligence (AI) Based Prediction of Mortality, ICU Admission and Ventilation Support Requirement for COVID-19 Patients Using 122 Clinical and Demographic Parameters"

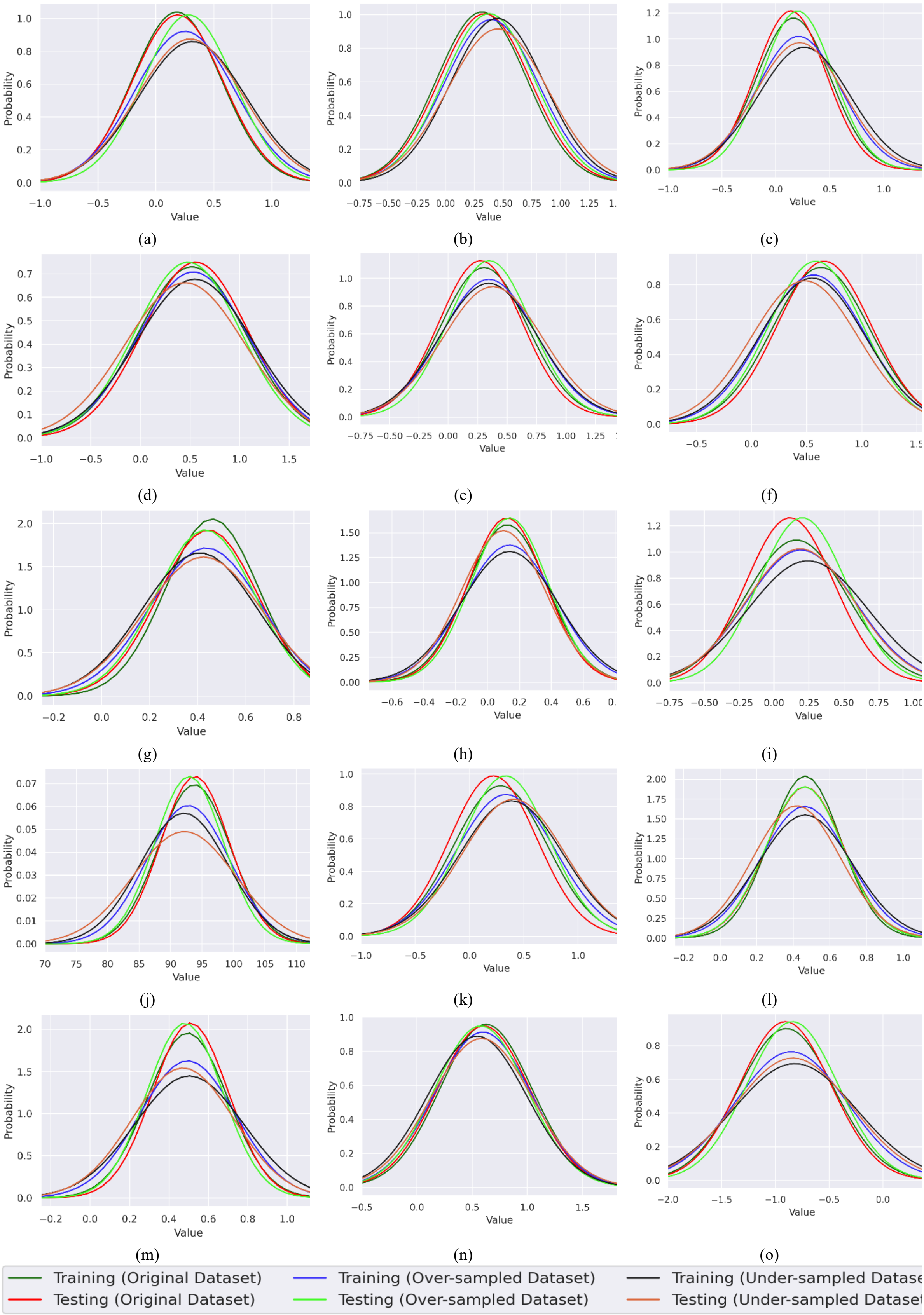


**Supplementary Figure 1:**
Probability distribution of training data and testing data for top features. (a) Acute kidney injury during hospitalization, (b)Age, (c) Therapeutic heparin, (d) Gender, (e) Troponin above 0.01, (f) Blood urea nitrogen above 20, (g) Blood pH above 7.45, (h) Acute hepatic injury during hospitalization (i) PulseOx

under 90, (j) Oxygen saturation in arterial blood by pulse oximetry, (k) Therapeutic exnox, (l) Blood pH between 7.35 and 7.45, (m) Blood pH below 7.35, (n) Procalcitonin below 0.25, (o) Kidney replacement therapy
